# Aerosol generation in children undergoing high flow nasal cannula therapy

**DOI:** 10.1101/2020.12.10.20245662

**Authors:** Elliott T. Gall, Aurélie Laguerre, Michelle Noelck, Annalise Van Meurs, Jared A Austin, Byron A. Foster

**Author notes:** **Address correspondence to:** Byron A. Foster, MD, MPH, 3181 SW Sam Jackson Park Rd, MC CDRC-P, Portland, OR 97239. **Financial disclosure:** The authors have indicated they have no financial relationships relevant to this article to disclose. **Data sharing statement:** Deidentified individual participant data will not be made available. Measurements of constituents in air made in participant rooms will be made available upon reasonable request. **Contributor statements:** Dr. Gall contributed to the study conception, design, data analysis and drafting the manuscript. Ms. Laguerre contributed to the study design, was the primary data collector, contributed to the analysis and writing the manuscript. Dr. Noelck contributed to the study design, execution of the study and revised the manuscript. Dr. Van Meurs contributed to the recruitment, data collection and revised the manuscript. Dr. Austin contributed to the study design and revised the manuscript. Dr. Foster contributed to the study conception, design, recruitment, data collection and drafting the manuscript.

## Abstract

**Objective:** High flow nasal cannula therapy (HFNC) may increase aerosol generation, putting health care workers at increased risk of infection, including from SARS-CoV-2. This study examined whether use of HFNC increases near-field aerosols and if there is a relationship between flow rate and near-field aerosol concentrations.

**Patients and Methods:** Subjects between 30 days and 2 years of age were enrolled. Each child received HFNC therapy at different flow rates over time. Three sampling stations with particle counters were deployed to measure aerosol generation and dispersion in the room: station one within 0.5 m of the subject, station two at 2 m, and station three on the other side of the room. We also measured carbon dioxide (CO_2_) and relative humidity. Station three (far-field) measurements were used to adjust the station one (near-field) measurements for room conditions.

**Results:** We enrolled ten children ranging from 6-24 months (median 9 months), two with respiratory illness. Elevated CO_2_ indicated the near-field measurements were in the breathing plane of the subjects. Near-field breathing plane concentrations of aerosols with diameter 0.3 – 10 µm are elevated by the presence of the patient with no HFNC flow, relative to the room far-field, by 0.45 #/cm^3^. While we observed variability between subjects in their emission and dispersion of particles, we did not find an association between HFNC and near-field elevations of particle counts.

**Conclusion:** Near-patient levels of particles with diameter in the 0.3-10 µm range was not affected by the use of HFNC in healthy patients. Further study on older children and children with increased mucus production may be warranted.

## Introduction

High flow nasal cannula therapy (HFNC) provides respiratory support for hospitalized children across a range of ages and diagnoses including asthma, pneumonia and bronchiolitis. World Health Organization (WHO) guidance suggests that HFNC does not cause wide-spread dispersion of droplets from patients.^1^ However, empirical data in clinical settings is lacking on whether HFNC contributes to aerosol generation. While children typically have more mild and even asymptomatic infections with SARS-CoV-2, respiratory disease and co-infection with other viruses have been reported.^2^ During the COVID-19 pandemic, HFNC has been treated as an aerosol generating procedure (AGP) in the United States given concern around particle generation, typically characterized in the health care field as transmission by droplet (≥5µm) and droplet nuclei (<5 µm).^3^ This requires use of N-95 masks, gowns, and other personal protective equipment when patients are receiving HFNC. Determining the risk of aerosol generation from HFNC has important implications for resource management and infection control measures.

Several studies have investigated transport of large droplets from patients undergoing HFNC. Kotoda et al.^4^ used a mannequin model to examine the effect of high flow nasal cannula at 60 L/min and observed large droplets (>50 µm) at 30 cm, but not 5 m from the mannequin’s face. A report examining adults coughing with and without the application of high flow nasal cannula (60 L/min) showed no significant difference in the distance of “visible” food-dye containing droplets;^5^ the length scale of droplets is not noted, though visible particles are often classified as those >100 um. These studies indicate large droplets are not effectively transported over long distances due to the forced air exiting the patient’s nasal and oral cavity

Studies have employed smoke as a tracer to evaluate impacts of HFNC on room air flows and as proxies of exhaled air exposure. Hui et al.^6^ used intrapulmonary smoke in a mannequin model to evaluate enhancement of exhaled air, measured by the extent of light-scattering as a function of distance from the patient. They showed an increase in “exhaled air dispersion” from 65 mm with HFNC flow of 10 L/min to 172 mm with HFNC flow of 60 L/min. Using smoke particles as tracers and an adult human head with a lung model attached, Elshof et al.^7^ examined the dispersion of 100 µm droplets using HFNC with a lung simulator. They described an estimated dispersion range of 100 µm droplets of between 18.8 and 33.4 cm from the individual using flow rates between 30-60 L/min. They also noted that HFNC increased the distance of exhaled smoke to nearly one meter under several conditions whereas a non-rebreather or Venturi mask did not influence the distance beyond normal breathing.^7^

This study sought to examine whether HFNC therapy use in children generates elevated particle levels in the near-field (0.5 m) of the patient’s breathing plane. We measured concentrations of particles and carbon dioxide (as an exhaled breath tracer) in rooms in a clinical care facility with varying HFNC flow rates for each patient. Our study addresses several knowledge gaps concerning HFNC and particle generation and dispersion as it: i) addresses an unstudied population, children, ii) was conducted in a clinical care facility with human subjects, and iii) directly measured aerosols with diameter 0.3 - 10 µm and carbon dioxide in the near-field breathing plane and room far-field. The goal was to generate data to inform the safe use of this therapy and inform resource management and infection control measures.

## Methods

This study was a prospective study looking at aerosol generation and dispersion from pediatric patients on high-flow nasal cannula (HFNC) in a typical pediatric hospital room.

### Subject eligibility and recruitment

Subjects were recruited through fliers and email announcements. Inclusion and exclusion criteria were gestation-corrected age 4 weeks to 24 months, no chronic cardiopulmonary conditions, and currently healthy with no Sars-Cov2 exposure or symptoms.

### Ethics

This study was approved as human subjects research through the OHSU IRB, and all parents provided written consent to participate.

### Experimental procedure

Hospital rooms were chosen from available pediatric acute care rooms (patient room, hereafter, and shown in Figure 1) and one procedure room at a tertiary care hospital in the Pacific Northwest. Patient and procedure rooms had floor area ∼24 m^2^ and ∼17 m^2^, respectively. CO_2_ tracer decay tests conducted in patient and procedure rooms resulted in an air-change rate of 8.4 and 11.0 h^-1^, respectively (Figure S1 of Supporting Information). Air entering the rooms is treated with MERV10 and MERV15 filtration.

**Figure 1.**
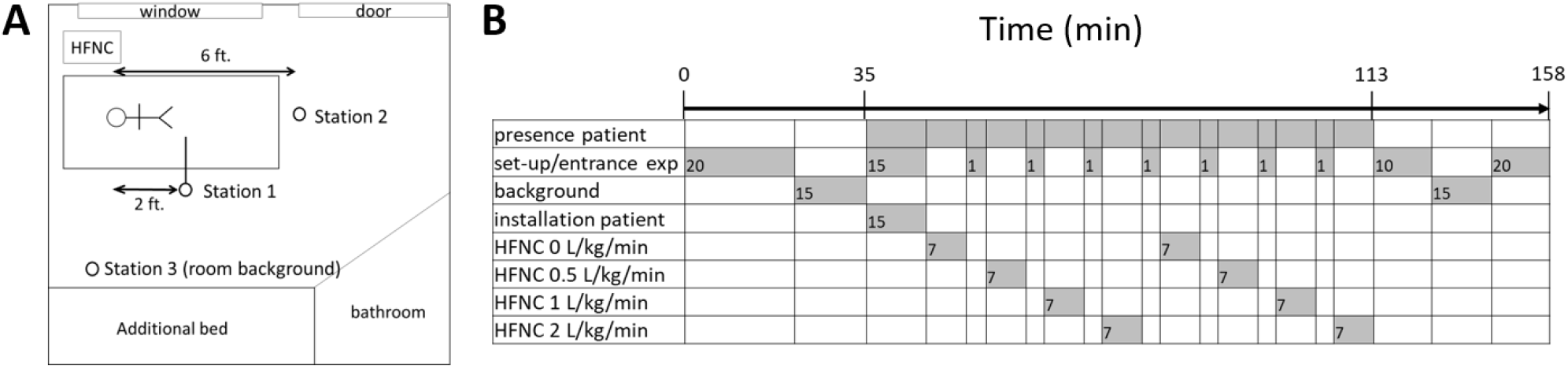
Panel A) Layout of patient room and sampling locations, and Panel B) Timeline of experiments for each patient

Subjects were placed on a hospital bed with a parent in the room, with the parent wearing a cloth or surgical mask at all times. A high-flow nasal cannula system (Fisher and Paykel’s Optiflow Junior, circuit RT 330) with an appropriately sized nasal cannula for each subject’s size and weight was set-up by a qualified respiratory therapist.

Ambient air in the hospital room was sampled with the door closed and no patient present (background condition) for 15 minutes. The child was then connected to the HFNC, flow was then increased from 0 to 0.5 L/kg/min, to 1 L/kg/min then finally to 2 L/kg/min, then back to 0 L/kg/min and repeated the cycle one more time for a total of two measurements per subject at each flow rate. Each cycle lasted about seven minutes. An experimental timeline is shown in Figure 1. HFNC air was heated to approximately 37°C and humidified. No supplementary oxygen was provided. We conducted a positive control following the completion of the protocol twice over the course of the study. In this control, particle and CO_2_ levels were measured in the breathing plane ∼0.5 m from the nasal/oral cavity of member of the research team during and after volitional coughing.

We recruited ten children ranging from 6-23 months (median nine months) and their parents to participate in the study between September and November 2020. The median weight of participants was 9.8 kg (range 7.3-14.0 kg). The flow rates were calculated for each child at 0.5 L/kg/min, 1 L/kg/min and 2 L/kg/min with a max flow rate in this study of 25 L/min, which two of the participants reached. Note that two patients (P02 and P03) were excluded from subsequent analysis as measurements occurred during periods of extremely elevated outdoor air pollution due to wildfires in the region; background particle concentrations were substantially elevated in the patient rooms. For patients with respiratory illness (P08 and P10), we were not able to vary the HFNC flowrate and could not access the room prior to patient occupancy for background measurement.

### Particle and carbon dioxide measurement

Three sampling stations were deployed in the room of each study participant prior to their arrival, excepting P08 and P10 who were present prior to sampling. The main sampling location (station 1, Figure 1) consisted of a common 0.9 m sampling line with inlet installed in the patient’s breathing plane at distance ∼0.5 m from the breathing zone, similar to O’Neil et. al^8^. The tubing was 0.95 cm outer diameter conductive tubing (Bev-a-line) with the inlet directed towards the patient. The sampling line was connected to a stainless-steel manifold with ports for four instruments. An optical particle sizer (TSI/OPS 3330) and scanning mobility particle sizer (TSI/NanoScan SMPS 3910) counted particles ranging from 0.01 to 10 μm at a time resolution of one-minute. A condensation particle counter (TSI, P-Trak 8525) measured particles ranging 0.02 to 1 μm in one second time interval. Isokinetic sampling was not possible due to the variability in airflows in the room and due to the exhalations of the patient. A CO_2_ analyzer (LICOR LI-820) measured CO_2_ levels in one second interval. A temperature and relative humidity sensor (Onset, S-THB-M002) measured in one-minute interval. The patient was not required to maintain a particular position to ensure their exhalations were directed towards the sampling inlet, per IRB requirements.

Two additional sampling stations (station 2 and 3, Figure 1) were installed to monitor the room. Each station included a low-cost particle counter (Purple Air, PA-II-SD), measuring particle number concentration in six size bins from 0.3 - 10 μm and recording every 80 seconds, and a CO_2_ sensor (Onset, MX1102) recording every minute. Sampling station 2 was installed ∼2 m from the patient’s breathing zone, roughly in line with the breathing plane established by sampling station 1. Sampling station 3 was placed on the other side of the room away from the patient. In this study, we normalize the data reported by station 1 (near-field) to that of station 3 (far-field), which we take as the ambient room particle and CO_2_ level. Note that we lacked particle number concentrations <0.3 μm in the station 2 and 3 locations; this investigation subsequently focuses on PM_0.3-10_.

### Field co-location of instruments

The Purple Air (PA) sensors and Onset CO_2_ sensors were co-located with the OPS and LICOR CO_2_ analyzer during a background period, where the room was unoccupied for 15 min. We used these periods to develop correction factors that were applied to the far-field (station 3) sensor during periods of participant occupancy. The OPS size bins were averaged to match the six bins of the PA. A correction factor for each size bin was calculated as in equation 1:

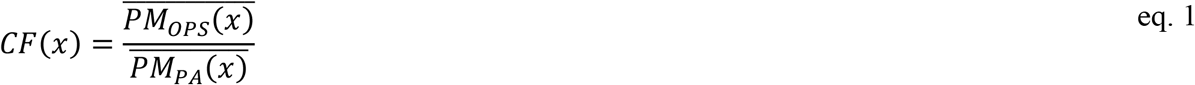

where *CF*(*x*) is the correction factor for size bin 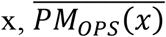 is the time-averaged OPS value in size bin x, and 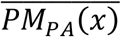 is the time-averaged PA value in size bin x.

For the CO_2_ sensors, we observed a greater range of background CO_2_ concentrations due to participants and researchers who were familiarizing and setting up the patient room, respectively. We used a linear regression to correct the values given by the fair-field CO_2_ sensor (Onset MX1102) to that of the LICOR LI-820 during the 15-minute background period.

### Calculations of ΔPM and ΔCO2

To account for the changing concentration of PM_0.3-10_ and CO_2_ in the room due to processes other than the patient undergoing HFNC, we normalize the near-field (station 1) measurements to that of the far-field (station 3). We report the normalized metrics as ΔPM_0.3-10_ (#/cm^3^) and ΔCO_2_ (ppm) calculated as shown in equations 2 and 3:

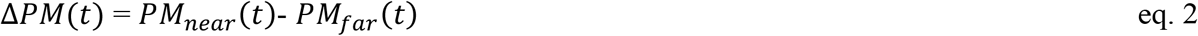

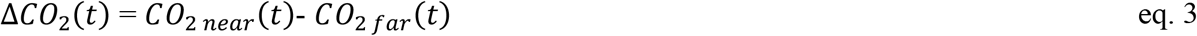

where *PM_near* (*t*) is the time-varying particle concentration at station 1 (#/cm^3^), *PM_far* (*t*)is the corrected (i.e., eq. 1) particle concentration at station 3, *CO*_(2 *near*) (*t*) is CO_2_ concentration at station 1 (ppm), and *CO*_2*far*_(*t*) is the corrected CO_2_ concentration at station 3 (ppm).

## Results

Measurements of particle concentrations, CO_2_, temperature and RH for two example patients are shown in Figure 2. In the top panel, room particle concentrations are reported in the near-field breathing plane (station 1) and far-field (station 3) of the room, with the second panel showing the difference (ΔPM_0.3-10_). For patient 01, near-field is generally higher than far-field, and coincided with generally positive, and during the second replicates sharp spikes in, measured ΔCO_2_. This implies measurements captured patient exhalations, as the only source of CO_2_ in the room is the patient. Conversely, in Patient 06 there are lower levels of particles in the near-field vs. far-field, possibly due to a low generation rate, room mixing conditions, and/or non-human sources of particles during this experiment. For positive controls we observe elevated particle and CO_2_ concentrations following the volitional cough.

**Figure 2.**
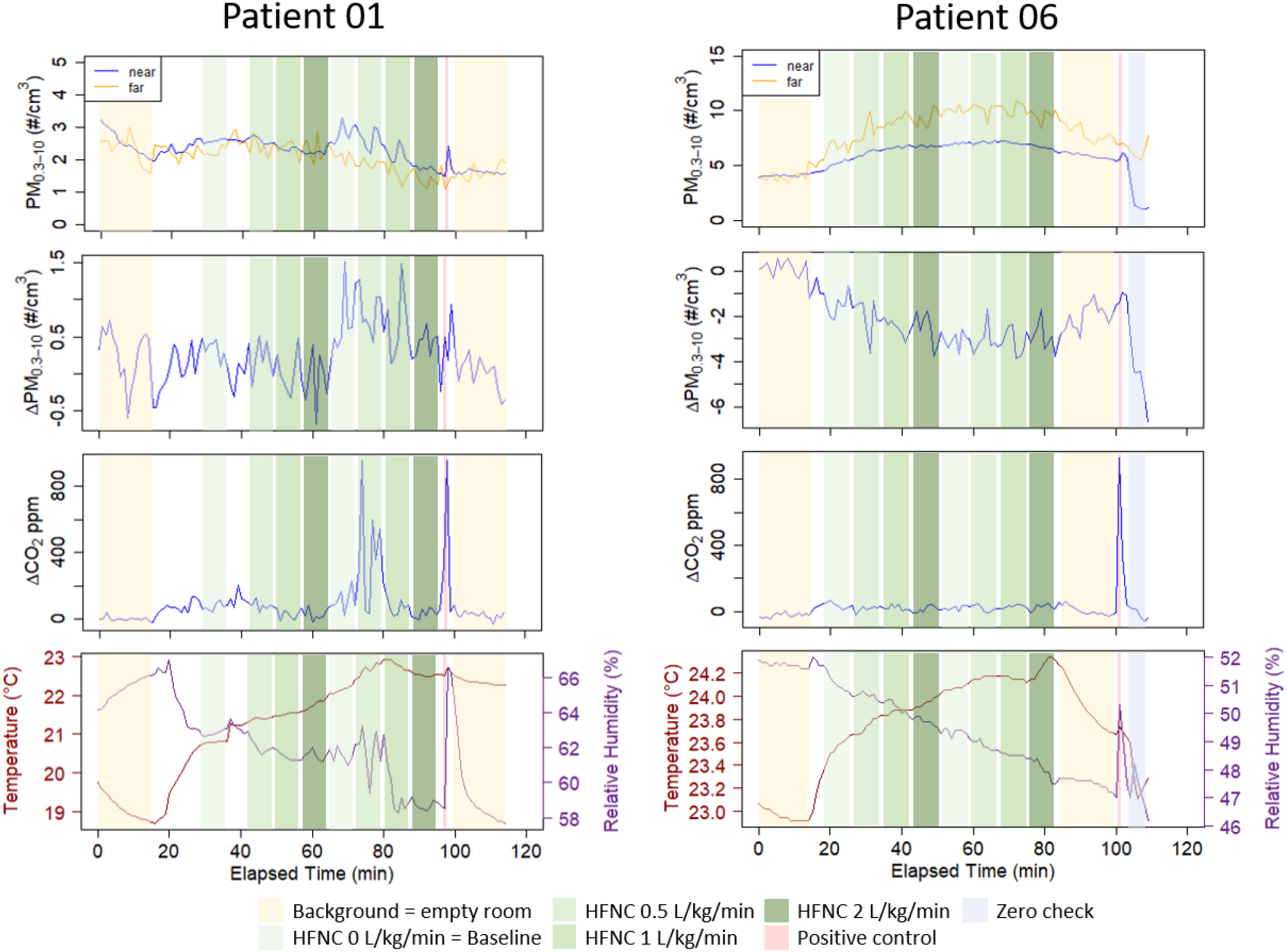
Example particle and CO_2_ concentrations in the breathing plane of two patients. Shading annotations shows the condition of the experimental protocol.

Across the six patients enrolled in the study with acceptable background PM, distributions of ΔPM and ΔCO_2_ are shown in Figure 3 across baseline conditions (HFNC at 0 L/kg/min), HFNC with flow, and positive control. Similar plots are shown for size-resolved particles in Figure S2 of the Supporting Information. Shown in Figure 3 are the 1-min measured raw data. Across all six patients, we observe that the presence of the patient alone (i.e., baseline) results in an increase in the near-field PM (i.e., median ΔPM_0.3-10_ is positive). Presence of HFNC flow does not significantly increase median ΔPM_0.3-10_ compared to the baseline condition.

**Figure 3.**
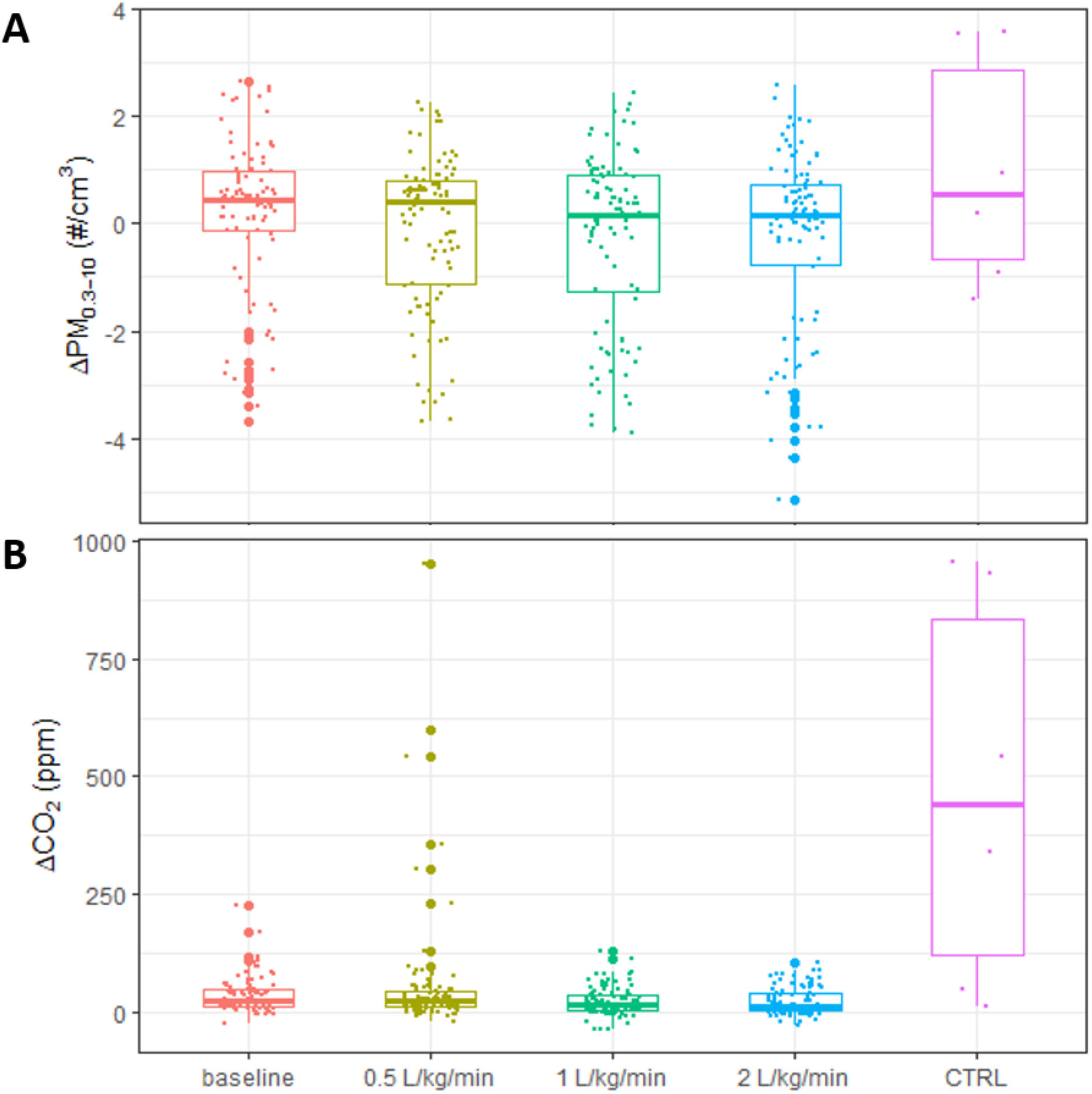
Panel A) Distributions of measured ΔPM_0.3-10_ and Panel B) ΔCO_2_ for six patients involved in this study. Centerline of box plots report median, extent of box is 25^th^ and 75^th^ percentiles, and whisker designates upper and lower extent of outliers in the distribution. Note that Δ indicates reported measurements are the difference between the near-field breathing plane and the coincident ambient room concentration (far-field), as explained in the text.

Measurements of ΔCO_2_ made for the six patients shown in Figure 3 indicate that median ΔCO_2_ is consistently positive. This implies that near-field measurements generally occurred in the exhalations of the patient. Again, no relationship is observed with HFNC flow. The volitional cough positive control resulted in substantially higher ΔCO_2_.

While median ΔPM_0.3-10 and_ ΔCO_2_ are consistently positive, there existed across-subject variability in ΔPM_0.3-10 and_ ΔCO_2._ For example, Patients 01, 05, 07, and 09 had consistently positive ΔPM_0.3-10_ while Patients 04 and 06 were consistently negative (Figure 4a). Values of ΔCO_2_ were more consistently positive than ΔPM_0.3-10_, as shown in Figure 4b, though again, there exists variability across subjects.

**Figure 4.**
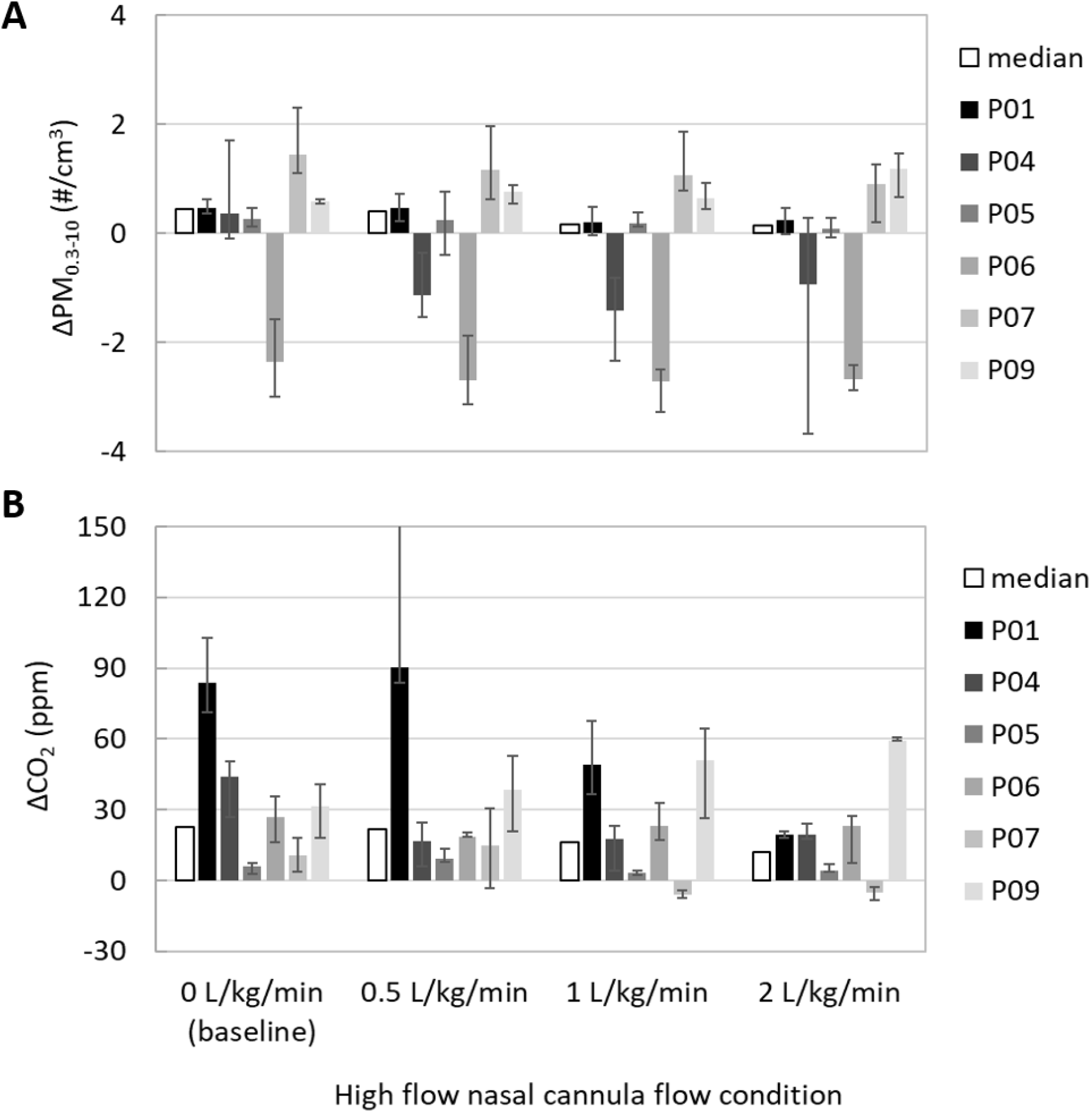
Panel A) Across-subjects variability in ΔPM_0.3-10_ and Panel B) ΔCO_2_. Each bar is the median across 1-min averaged measurements at each HFNC flow condition for the indicated subject. The error bars show the range across the 1-min averaged measurements (max-min). The upper error bar for P01 at 0.5 L/kg/min extends to 310 ppm, not shown for figure clarity.

In Figure 5, we show the results of ΔPM_0.3-10 and_ ΔCO_2_ for the two patients recruited who had respiratory illness; results are limited to only one flowrate as we did not alter the patients’ care directives. Since background measurements were infeasible, correction factors were used from healthy patient studies conducted on the same respective days. As in healthy patients, we observe variability in median ΔPM_0.3-10_, with P08 negative and P10 positive. In contrast, ΔCO_2_ for both patients is greater than zero, implying measurements occurred in patient breathing planes.

**Figure 5.**
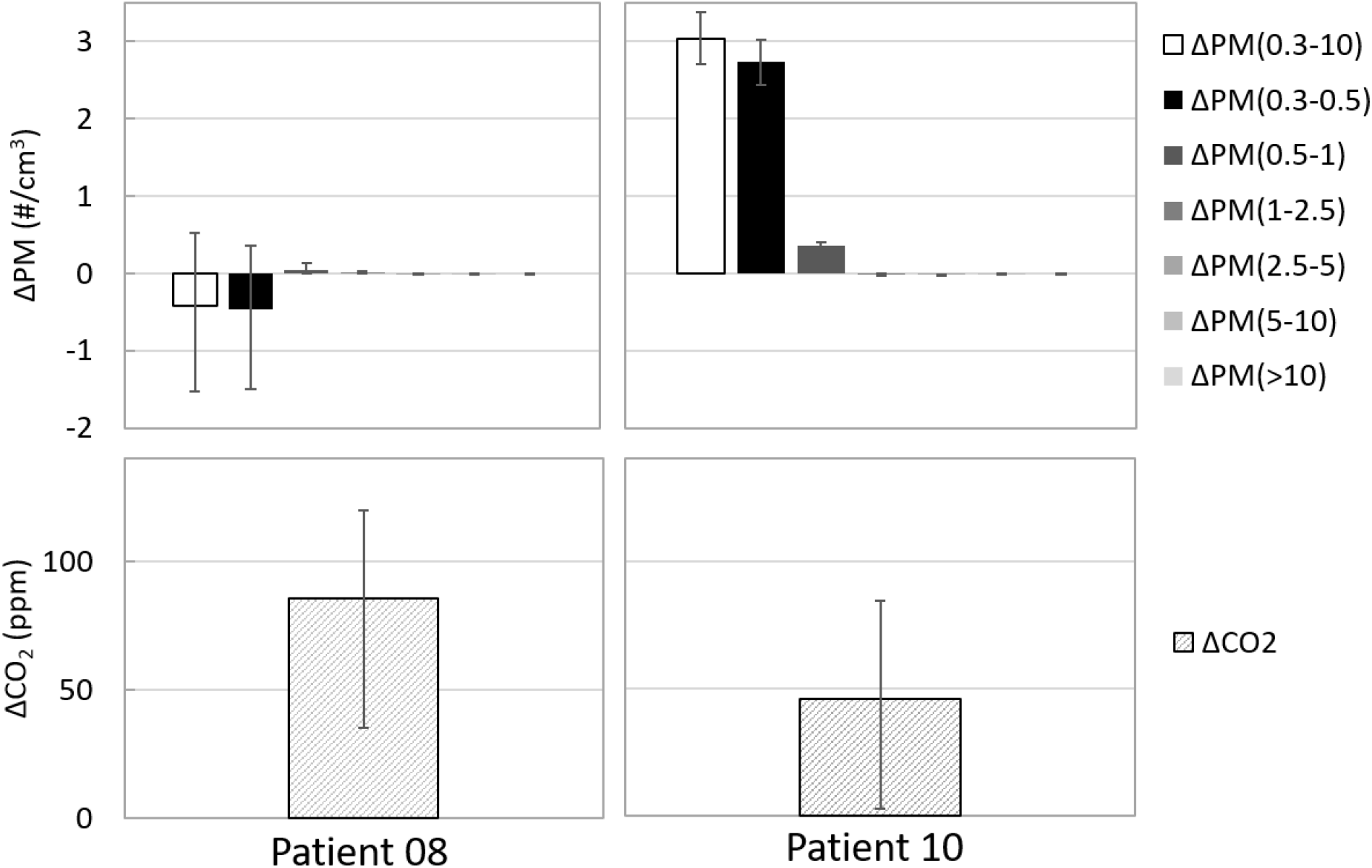
Size resolved ΔPM_0.3-10_ and ΔCO_2_ for two patients with diagnosed respiratory illness. Patient 08 was 3 months old and HFNC flowrate of 3 LPM, Patient 10 was 24 months with HFNC flowrate of 15 LPM. Bars show median values of 1-min averaged measurements while error bars show the range across a 10-min monitoring period.

## Discussion

Results of this study indicate, across patients, that HFNC is not a substantial source of aerosol generation in the near-field beyond that of the patient’s presence. Human breath contains particles; while results are variable across time for each patient and across patients, the median ΔPM_0.3-10_ reported in this measurement is roughly consistent with the previous measurements of particle number concentrations on human breath. Johnson et al.^9^ report particle levels in speaking and coughing emissions in the size range of 0.5 - 1000 µm of 0.16 #/cm^3^ and 0.22 #/cm^3^, respectively. In this study, the complex fluid mechanics occurring in the patient’s breathing plane due to exhaled breath, HFNC airflow, and the room airflows complicate further theoretical calculations of particle concentrations or emission rate originating from the patient. Humans also generate particles from activity,^10^ particles originating from the respiratory system versus, e.g., patient movement, cannot be differentiated here.

Median values of ΔPM_0.3-10_ decreased slightly, though not statistically significantly, with increasing HFNC flow rate. We speculate this may be the result of enhanced mixing between forced air from subject and room air with higher velocities at higher HFNC flow conditions. We evaluate statistical significance of differences in medians of ΔPM_0.3-10_ and ΔCO_2_ across HFNC flow rates using a Wilcoxon rank sum test for ΔPM_0.3-10_ and a student t-test for ΔCO_2_, based on a Kolmogorv-Smirnov test for normality. Tests for normality and statistical testing employed the average of each HFNC condition conducted in duplicate across the six subjects (i.e., 12 independent samples of ΔPM_0.3-10_ and ΔCO_2_ for each condition). There are no statistically significant differences across ΔPM_0.3-10_ or ΔCO_2_ for any comparison of flow conditions. We set the threshold of significance as *p* < 0.0083 for 95% confidence with Bonferonni correction for multiple comparisons. Calculated *p*-values are shown in the Table S1 of the Supporting Information.

Results shown in Figure 4 reveal high variability in near-patient concentrations of PM and CO_2_. The explanation for the mechanism behind these observations is beyond the scope of this paper, though we speculate it is possible that patients with negative ΔPM_0.3-10_ may be low emitters of particles or positioned in the space such that enhanced particle deposition is occurring in the turbulence generated from airflows interacting with the patient and associated equipment (bedding, instruments, etc.). Particles also deposit in the respiratory system.^11^ Patient 06 and Patient 04 measurements were conducted during relatively high room background PM levels, perhaps contributing to the negative ΔPM_0.3-10_ observed. We note that prior studies have observed large variability in particle emission rate and concentrations in exhalations of humans during breathing and speaking.^12–16^ There is debate on the size of particles that are considered infectious, with droplet nuclei playing a larger role than previously considered^17^ – our study measured a broad range of potentially infectious particles including droplet nuclei.

Differences in near-field to far-field CO_2_ were larger and more pronounced than for PM. CO_2_ levels in human breath are ∼100x higher than ambient levels (∼38,000 vs. 400 ppm).^18^ In contrast, particle concentrations in human breath in the size range 0.3 - 10 um are expected to be similar or lower than background levels measured in the patient rooms, e.g., Fairchild and Stampfer^12^ report particles in exhaled breath of <0.1 to 4 #/cm^3^. Notably, there is wide variation in particle concentrations in human breath and particle generation rates during coughing, with the presence of a respiratory infection and causing increased particle generation rate.^19^

In contrast to the variability in ΔPM_0.3-10_ shown, ΔCO_2_ is variable but more consistently positive (Figure 4b), implying that measurements were generally made in the breathing planes of the patients. There does not appear to be a relationship between elevated ΔCO_2_ and ΔPM_0.3-10_, that is, high values of ΔCO_2_ do not necessarily associate with high ΔPM_0.3-10_. For example, P01 had the highest ΔCO_2_ for three of four HFNC flow conditions, but ΔPM_0.3-10_ was consistently near the median value reported. Again, we speculate that this is a result of differences in particle generation across subjects that are not related to metabolism (e.g., unknown physiological factors that have been previously suggested as explaining “superemission” of aerosol during speech^13^).

Our limited sample of two patients with respiratory illness shown in Figure 5 demonstrates variability in near-field elevations of particles, with Patient 10 showing greater ΔPM_0.3-10_ than all healthy patients by a substantial margin. This appears largely driven by a difference in the behavior of particles 0.3 – 0.5 µm, as the smallest size bin measured this size range dominates the particle number concentration. For both patients with respiratory infection we note there was an elevation in ΔPM_0.5-1_, a size range that a prior study shows is statistically significantly elevated during a respiratory infection.^20^ However, we did not have the ability to vary HFNC flowrate, and so lack a baseline period of no HFNC flow for comparison.

## Conclusions

Our measurements indicate near-field (∼0.5 m) breathing plane concentrations of aerosol and carbon dioxide are elevated by the presence of the patient with no HFNC flow, relative to the room far-field, by 0.45 #/cm^3^ and 23 ppm, for PM and CO_2_, respectively. Addition of HFNC flow in the range of 0.5 - 2 L/kg/min does not statistically significantly change the magnitude of near-field PM or CO_2_, corrected for the room far-field. These findings indicate that HFNC use in children may not substantially elevate clinician aerosol exposures greater than the presence of the patient alone, though we observe variability across patients. However, we caution that most of our empirical data were collected for healthy patients – our limited data for HFNC in children with respiratory illness showed one patient with substantially elevated near-field ΔPM_0.3-10_ while for another patient we observe a small decrease in this metric. Further study of the impacts of HFNC on particle generation and dispersion in patients with respiratory illness is warranted.

## Data Availability

Deidentified individual participant data will not be made available. Measurements of constituents in air made in participant rooms will be made available upon reasonable request.

## Acknowledgements

We thank the families who volunteered to participate in the study. Paula Bennett, MSN, MHA, RN, Brenda Rickert, MSN, RN, Emily Palmquist, MSN, RN, Rachel Coleman, BS, RRT-NPS, many respiratory therapy staff and the Hospital Chief Engineer, Andrew Wilkes, contributed to the completion of this study. We also thank Georgina Bicknell, MS, RN and Dana Braner, MD for their support.

## Notes

**Funding source:** The Doernbecher Philanthropy Board provided funding for the study.

**Conflict of interest statement:** The authors have indicated they have no potential conflicts of interest to disclose.

### Competing Interest Statement

The authors have declared no competing interest.

### Funding Statement

The Doernbecher Philanthropy Board provided funding for the study.

### Author Declarations

The Oregon Health & Science University IRB approved this study as human subjects research. All parents provided written consent to participate.

## References

1. WHO. Clinical Management of Severe Acute Respiratory Infection When Novel Coronavirus (2019-NCoV) Infection Is Suspected. World Healthy Organization; 2020. Accessed December 5, 2020. https://www.who.int/publications-detail-redirect/clinical-management-of-covid-19

2. Wu Q, Xing Y, Shi L, et al. Coinfection and Other Clinical Characteristics of COVID-19 in Children. Pediatrics. 2020;146(1). doi:10.1542/peds.2020-0961

3. Agarwal A, Basmaji J, Muttalib F, et al. High-flow nasal cannula for acute hypoxemic respiratory failure in patients with COVID-19: systematic reviews of effectiveness and its risks of aerosolization, dispersion, and infection transmission. Can J Anaesth. Published online June 15, 2020:1–32. doi:10.1007/s12630-020-01740-2

4. Kotoda M, Hishiyama S, Mitsui K, et al. Assessment of the potential for pathogen dispersal during high-flow nasal therapy. J Hosp Infect. 2020;104(4):534–537. doi:10.1016/j.jhin.2019.11.010

5. Loh N-HW, Tan Y, Taculod J, et al. The impact of high-flow nasal cannula (HFNC) on coughing distance: implications on its use during the novel coronavirus disease outbreak. Can J Anaesth. 2020;67(7):893–894. doi:10.1007/s12630-020-01634-3

6. Hui DS, Chow BK, Lo T, et al. Exhaled air dispersion during high-flow nasal cannula therapy versus CPAP via different masks. Eur Respir J. 2019;53(4). doi:10.1183/13993003.02339-2018

7. Elshof J, Hebbink RHJ, Duiverman ML, Hagmeijer R. High-flow nasal cannula for COVID-19 patients: risk of bio-aerosol dispersion. Eur Respir J. 2020;56(4). doi:10.1183/13993003.03004-2020

8. O’Neil CA, Li J, Leavey A, et al. Characterization of Aerosols Generated During Patient Care Activities. Clin Infect Dis. 2017;65(8):1335–1341. doi:10.1093/cid/cix535

9. Johnson GR, Morawska L, Ristovski ZD, et al. Modality of human expired aerosol size distributions. Journal of Aerosol Science. 2011;42(12):839–851. doi:10.1016/j.jaerosci.2011.07.009

10. Ferro AR, Kopperud RJ, Hildemann LM. Source strengths for indoor human activities that resuspend particulate matter. Environ Sci Technol. 2004;38(6):1759–1764. doi:10.1021/es0263893

11. Hinds WC. Aerosol Technology: Properties, Behavior, and Measurement of Airborne Particles. 2 edition. Wiley-Interscience; 1999.

12. Fairchild CI, Stampfer JF. Particle Concentration in Exhaled Breath. American Industrial Hygiene Association Journal. 1987;48(11):948–949. doi:10.1080/15298668791385868

13. Asadi S, Wexler AS, Cappa CD, Barreda S, Bouvier NM, Ristenpart WD. Aerosol emission and superemission during human speech increase with voice loudness. Scientific Reports. 2019;9(1):2348. doi:10.1038/s41598-019-38808-z

14. Edwards DA, Man JC, Brand P, et al. Inhaling to mitigate exhaled bioaerosols. PNAS. 2004;101(50):17383–17388. doi:10.1073/pnas.0408159101

15. Papineni RS, Rosenthal FS. The Size Distribution of Droplets in the Exhaled Breath of Healthy Human Subjects. Journal of Aerosol Medicine. 1997;10(2):105–116. doi:10.1089/jam.1997.10.105

16. Fabian P, McDevitt JJ, DeHaan WH, et al. Influenza Virus in Human Exhaled Breath: An Observational Study. PLOS ONE. 2008;3(7):e2691. doi:10.1371/journal.pone.0002691

17. Fennelly KP. Particle sizes of infectious aerosols: implications for infection control. The Lancet Respiratory Medicine. 2020;8(9):914–924. doi:10.1016/S2213-2600(20)30323-4

18. Rudnick SN, Milton DK. Risk of indoor airborne infection transmission estimated from carbon dioxide concentration. Indoor Air. 2003;13(3):237–245.

19. Lindsley WG, Pearce TA, Hudnall JB, et al. Quantity and Size Distribution of Cough-Generated Aerosol Particles Produced by Influenza Patients During and After Illness. J Occup Environ Hyg. 2012;9(7):443–449. doi:10.1080/15459624.2012.684582

20. Lee J, Yoo D, Ryu S, et al. Quantity, Size Distribution, and Characteristics of Cough-generated Aerosol Produced by Patients with an Upper Respiratory Tract Infection. Aerosol Air Qual Res. 2019;19(4):840–853. doi:10.4209/aaqr.2018.01.0031

