## Supplemental Data for "Aerosol generation in children undergoing high flow nasal cannula therapy"

**Supporting Information for: Aerosol generation in children undergoing high flow nasal cannula therapy**

**Number of Pages: 4**

**Number of Tables: 1**

**Number of Figures: 2**

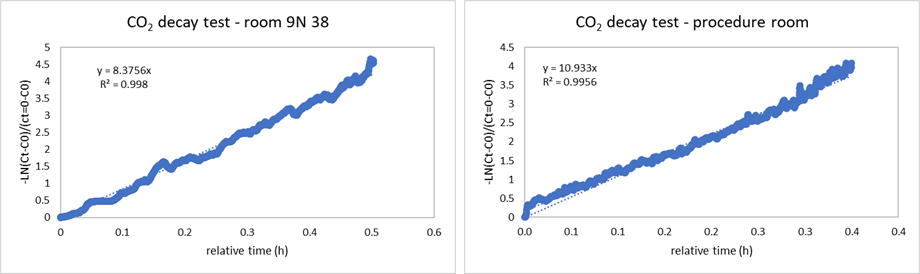

**Figure S1**. Results of tracer decay tests to estimate air change rate through the “patient room”, labeled as room 9N38 and the procedure room.

**Instrument calibration**

The OPS was calibrated by the manufacturer two weeks before the beginning of the experiments. The SMPS was calibrated by the manufacturer less than a year before the beginning of the experiments. The LICOR 820 was calibrated at different concentrations using a pure CO_2_ gas cylinder in our laboratory.

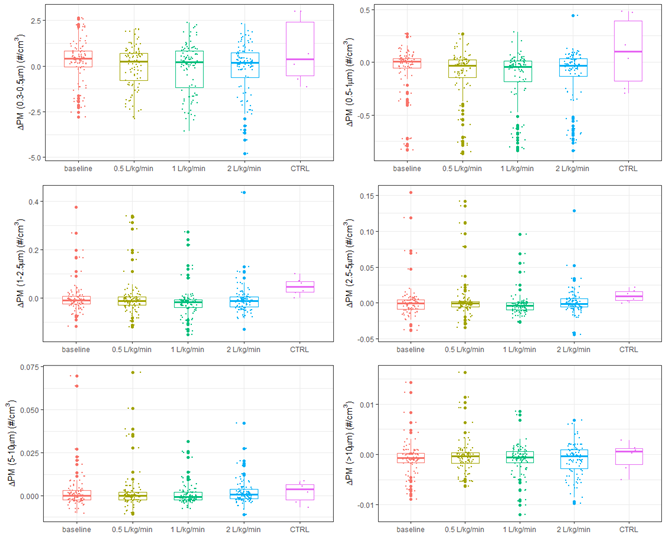

**Figure S2**. Summary of $\Delta$PM measurements disaggregated into six size bins.

**Table S1**. Summary of p-values from comparison of $\Delta$PM_0.3-10_ and $\Delta$CO_2_ across flow conditions.

|  | BL-0.5* | BL-1.0 | BL-2.0 | 0.5-1 | 0.5-2 | 1-2 |
| --- | --- | --- | --- | --- | --- | --- |
| $\Delta$PM_0.3-10_ | 1 | 0.3408 | 0.3708 | 0.5067 | 0.3708 | 0.8399 |
| $\Delta$CO_2_ | 0.1689 | 0.0959 | 0.1949 | 0.0794 | 0.2974 | 0.8322 |

*BL = baseline while numbers refer to HFNC flowrate in units of L/min/kg
